# CT-based Automated Volumetry as a Biomarker of Global and Split Renal Function in Living Kidney Donors

**DOI:** 10.64898/2026.02.24.26346974

**Authors:** Anna Fink, Franziska Burzer, Vlad Sacalean, Stephan Rau, Kai Kästingschäfer, Alexander Rau, Anna Köttgen, Fabian Bamberg, Bernd Jänigen, Maximilian F Russe

## Abstract

**Background:** Kidney volumetry derived from CT has been proposed as a surrogate of renal function in living kidney donor evaluation. However, clinical integration has been limited by reader-dependent workflows and semiautomatic methods susceptible to image quality.

**Purpose:** To evaluate whether fully automated CT-based segmentation of renal cortex, medulla and total parenchymal volume provides reproducible volumetric biomarkers associated with global and split renal function in living kidney donor candidates.

**Materials and Methods:** In this retrospective single-center study, 461 living kidney donor candidates (2003–2021) underwent contrast-enhanced abdominal CT. A convolutional neural network was trained to automatically segment cortical, medullary, and total parenchymal volumes on arterial-phase images. Segmentation performance was evaluated against manual reference annotations. Volumes were indexed to body surface area. Associations with eGFR, 24-hour creatinine clearance, cystatin C, and tubular clearance were assessed using Spearman correlation coefficient (ρ), and side-specific volume fractions were compared with scintigraphy -derived split function.

**Results:** Automated segmentation achieved excellent agreement with expert reference segmentations (Dice 0.95 for cortex; 0.90 for medulla). eGFR correlated moderately with cortical (ρ = 0.46) and total parenchymal volume (ρ = 0.45), and modestly with medullary volume (ρ = 0.30). Similar associations were observed for other global measures, with the strongest correlation for cortical volume and tubular clearance (ρ = 0.53). Side-specific volume fractions correlated with scintigraphy-derived split renal function (ρ = 0.49–0.56; all p < 0.001).

**Conclusion:** Automated CT-based renal subcompartment segmentation provides reproducible volumetric biomarkers within routine donor evaluation. Cortical volume performs comparably to total parenchymal volume and tracks split renal function at the cohort level, suggesting potential utility in donor assessment.

## Introduction

As the demand for kidney transplantation continues to exceed organ availability, living donor kidney transplantation remains a central strategy to expand access while preserving excellent recipient outcomes (1). Accurate donor evaluation is essential and typically integrates global renal function with assessment of relative kidney contribution, particularly when asymmetry or anatomic complexity is present, to guide graft-side selection and protect donor renal reserve (2).

Multiphase CT is routinely performed in donor candidates for vascular mapping and anatomic assessment, enabling quantitative biomarker extraction from data already embedded in clinical workflows (2). In living donor cohorts, CT-derived total kidney volume has shown clinically meaningful associations with renal function measures and post-donation functional outcomes, suggesting that morphologic metrics reflect functional renal mass (3,4).

However, total kidney volume is a composite metric that does not resolve biologically distinct renal compartments. Histopathologic analyses of donor kidneys link lower cortical volume and reduced cortex-to-medulla ratio to nephrosclerosis, whereas higher cortical volume reflects larger glomerular structures, supporting a structural basis for functional variability (5). Consistent with this, imaging studies have demonstrated that cortical volume correlates with glomerular filtration (6,7).

To date, subcompartment analyses have largely relied on manual or semi-automated segmentation approaches that are time-intensive and difficult to standardize (8). This methodological variability contributes to heterogeneity across studies and limits translation into routine donor workflows (8). Recent advances in deep learning-based segmentation have made the automated extraction of renal substructures technically feasible (9), providing the foundation for a fully automated compartment-resolved volumetry pipeline.

Leveraging this approach, we trained and implemented a fully automated arterial-phase CT segmentation network in a large cohort of living kidney donor candidates to (i) quantify associations between cortical, medullary, and total parenchymal volumes and established pre-donation functional metrics and (ii) compare side-specific volume fractions with scintigraphy-derived split renal function.

## Materials and Methods

### Study Design and Population

This retrospective single-center study included consecutive living kidney donor candidates who underwent multiphase abdominal CT between January 2003 and December 2021 as part of routine donor evaluation. The institutional ethics committee approved the study (Ethics Committee University of Freiburg, EK: 22-1270-S1-retro) and waived written informed consent.

Eligible candidates were required to have (i) an arterial-phase CT suitable for automated renal segmentation and (ii) at least one available pre-donation renal function metric, including serum creatinine-based estimated glomerular filtration rate (eGFR), 24-hour urinary creatinine clearance, serum cystatin C, and/or scintigraphy-derived tubular clearance or split renal function.

Exclusion criteria included severe motion artifacts, incomplete arterial-phase kidney coverage, and corrupted or incomplete imaging data.

### CT and Scintigraphy Acquisition

CT examinations were performed using a standardized multiphase abdominal protocol for vascular mapping and anatomic assessment. The protocol included arterial, portal venous, and excretory phases. Arterial-phase acquisition was initiated using bolus-tracking technology. Examinations were performed on multidetector CT scanners from Siemens Healthineers and Toshiba Medical Systems, with a small proportion from Agfa HealthCare systems. Images were reconstructed with vendor-specific soft-tissue kernels at 1.0-mm or 0.5-mm slice thickness.

Renal scintigraphy was performed in a subset of 452 candidates as part of routine donor evaluation using 99mTc-mercaptoacetyltriglycine (MAG3) in accordance with institutional protocols.

### Automated Kidney Subcompartment Segmentation

Bilateral renal cortex and medulla were manually segmented on 50 arterial-phase donor CT scans by a radiology resident (4 years of experience) to create a training dataset. Ten percent of cases were randomly withheld for internal validation during model development.

A Deep Neural Patchwork-based three-dimensional convolutional neural network was trained for multi-scale segmentation of cortex and medulla using nested 3D patches (10). The architecture combined coarse contextual information with high-resolution refinement and was trained with weighted cross-entropy loss and Adam optimization. Data augmentation included random rotations and isotropic scaling. Implementation was performed in Python within the NORA framework (11) and trained on an NVIDIA RTX A6000 GPU.

The trained network was subsequently applied to the remaining cohort to generate separate masks for left and right cortex and medulla. Total parenchymal volume was defined as the sum of cortical and medullary volumes. The renal hilum, collecting system, and parenchymal cysts were excluded by model definition. All automated segmentations underwent structured visual quality control by a radiologist. Cases with incomplete coverage or gross segmentation errors were excluded from volumetric analyses.

Interobserver variability was assessed in a subset of 20 cases independently segmented by a second radiology resident (2 years of experience), with Dice similarity coefficients calculated between readers.

Volumes were calculated as voxel count multiplied by voxel volume derived from DICOM metadata. For primary analyses, volumes were normalized to body surface area (BSA, mL/m²), calculated using the Mosteller formula (12) based on height and weight obtained from DICOM metadata. For split-function analyses, the relative volume contribution of each kidney was calculated as:

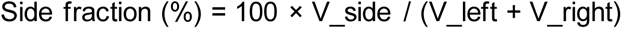

### Functional metrics

All renal function measures were obtained as part of the standard pretransplant evaluation and were recorded in temporal proximity to CT. Global renal function was assessed using serum creatinine-based eGFR (ml/min/1.73m^2^) calculated with the 2021 CKD-EPI equation (13). When available, 24-hour urinary creatinine clearance (mL/min) and serum cystatin C (mg/L) were extracted from clinical records. Renal scintigraphy provided tubular clearance (mL/min/1.73 m2) and relative split renal function expressed as the percentage contribution of each kidney.

### Statistical Analysis

Associations between CT-derived renal volumes and global renal function metrics were assessed using Spearman rank correlation coefficients (ρ), given potential non-normal distributions. Primary analyses used BSA-normalized renal volumes. Correlations were calculated with indexed eGFR (mL/min/1.73 m²) as well as with non-indexed laboratory measures, including cystatin C and 24-hour creatinine clearance. As a sensitivity analysis to evaluate potential effects of body-size indexing, correlations were additionally computed between absolute renal volumes and absolute eGFR (mL/min).

For split renal function, Spearman correlations were calculated between scintigraphy - derived split function (%) and CT-derived volume fractions for cortex, medulla, and total parenchymal volume. Agreement between CT-derived and scintigraphy-derived split renal function was evaluated using Bland-Altman analysis. Differences between CT-derived and scintigraphy-derived volume fractions were plotted against their mean. Mean bias and 95% limits of agreement (mean difference ± 1.96 × standard deviation) were calculated.

Clinically relevant split asymmetry was defined as an absolute left-right difference ≥10 percentage points on scintigraphy (approximately corresponding to a 55/45 distribution). Discriminative performance of CT-derived asymmetry was assessed using receiver operating characteristic (ROC) analysis, with area under the curve (AUC) reported for cortical, medullary, and total parenchymal volume fractions.

Statistical analyses were performed in Python (version 3.12.12) using pandas (2.2.2), NumPy (2.0.2), SciPy (1.16.3), statsmodels (0.14.6), and scikit-learn (1.6.1). A two-sided p value < 0.05 was considered statistically significant.

## Results

### Cohort Characteristics and Segmentation Performance

During the study period, 537 living kidney donor candidates underwent multiphase CT. Twenty-four were excluded due to severe motion artifacts (n = 5), incomplete arterial-phase coverage (n = 16), or corrupted imaging data (n = 3), leaving 513 examinations suitable for automated segmentation (Figure 1).

**Figure 1.**
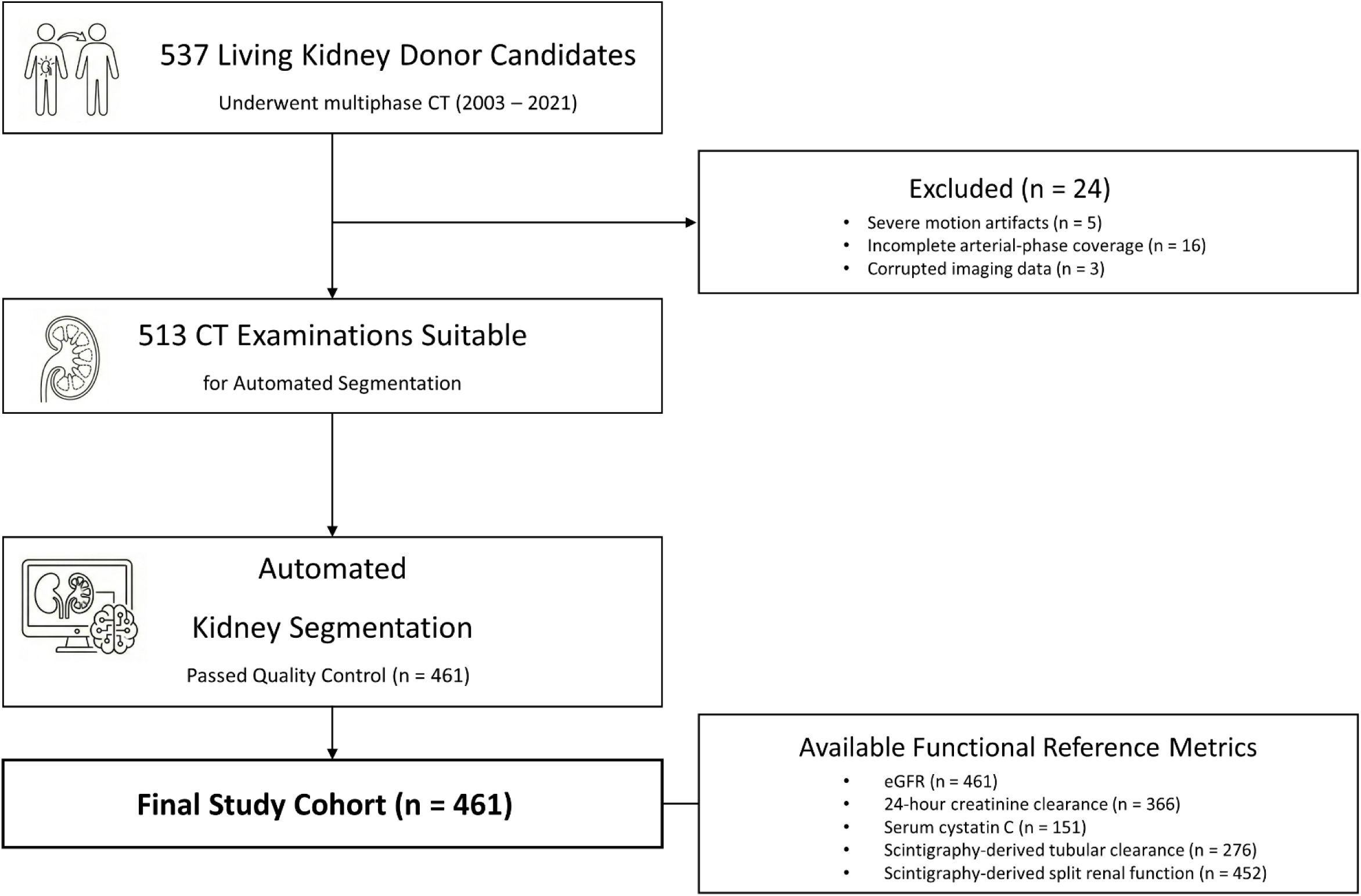
Study cohort flow diagram. Of 537 living kidney donor candidates who underwent multiphase CT, 24 were excluded, with 461 cases in the final analytic cohort after automated segmentation quality control. Availability of functional reference measures varied across endpoints.

The cohort had a mean age of 51.4 ± 9.5 years, 61.8% were female, and mean eGFR was 96.3 ± 12.4 mL/min/1.73 m². Additional functional reference measures were available in subsets, including 24-hour creatinine clearance (n = 366) and serum cystatin C (n = 151). Renal scintigraphy was performed in 452 subjects, providing tubular clearance (n = 276) and split renal function (n = 452). Detailed information is shown in Table 1.

**Table 1.**
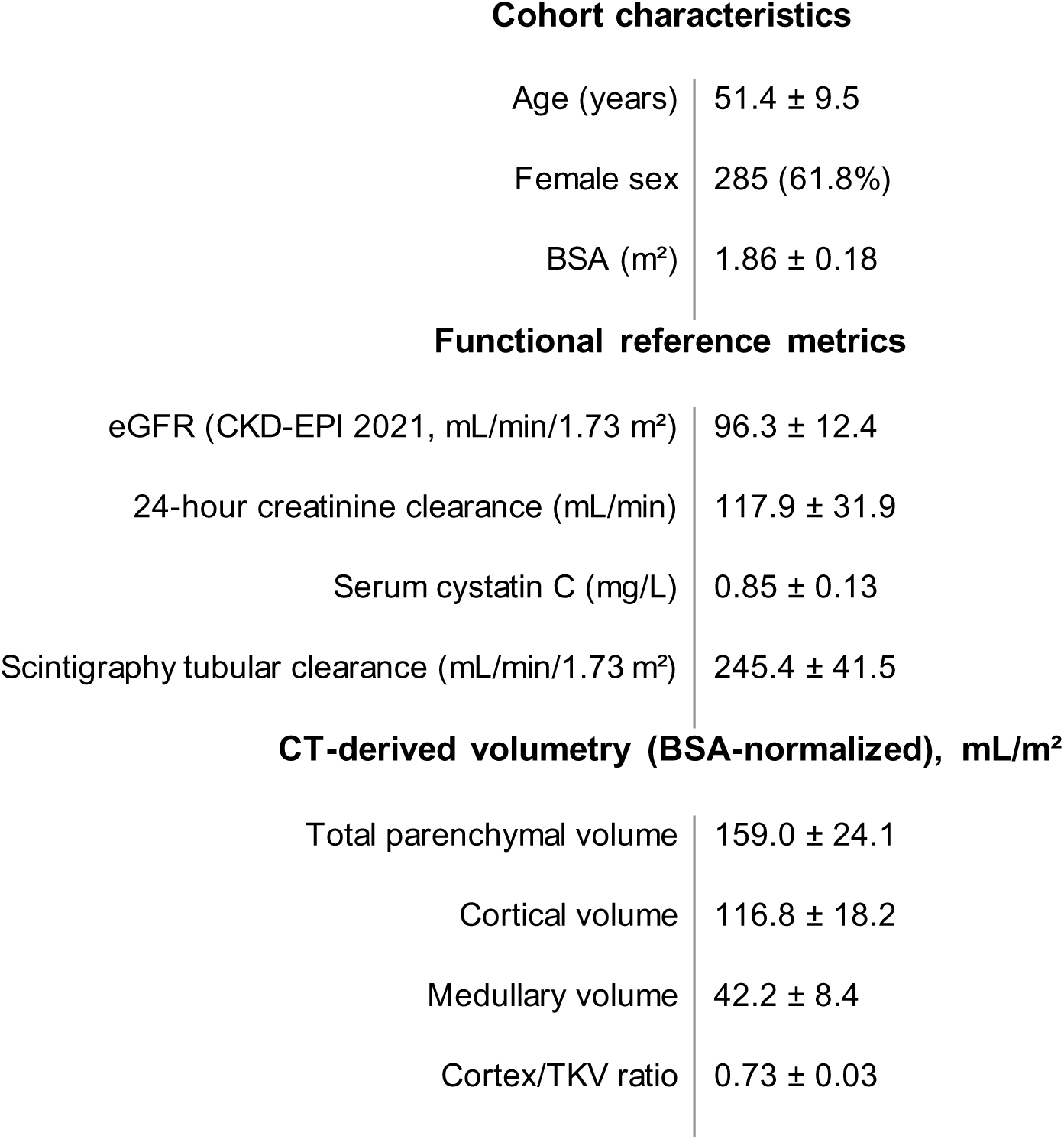
Cohort characteristics, functional reference metrics, and CT-derived renal volumes. Data are mean ± standard deviation unless otherwise indicated. Split renal function and tubular clearance were available in subsets based on clinical practice. BSA = body surface area.

| Cohort characteristics |  |
| --- | --- |
| Age (years) | 51.4 ± 9.5 |
| Female sex | 285 (61.8%) |
| BSA (m<sup>2</sup>) | 1.86 ± 0.18 |
| Functional reference metrics |  |
| eGFR (CKD-EPI 2021, mL/min/1.73 m<sup>2</sup>) | 96.3 ± 12.4 |
| 24-hour creatinine clearance (mL/min) | 117.9 ± 31.9 |
| Serum cystatin C (mg/L) | 0.85 ± 0.13 |
| Scintigraphy tubular clearance (mL/min/1.73 m<sup>2</sup>) | 245.4 ± 41.5 |
| CT-derived volumetry (BSA-normalized), mL/m<sup>2</sup> |  |
| Total parenchymal volume | 159.0 ± 24.1 |
| Cortical volume | 116.8 ± 18.2 |
| Medullary volume | 42.2 ± 8.4 |
| Cortex/TKV ratio | 0.73 ± 0.03 |

CT examinations were performed on Toshiba Medical Systems (50.5%), Siemens Healthineers (47.3%), and Agfa HealthCare (2.2%) scanners. Tube voltage ranged from 70 to 140 kVp and was 120 kVp in the majority of examinations (96.8%). Reconstruction slice thickness was 1.0 mm in 94.1% and 0.5 mm in 5.9% of examinations.

Automated segmentation demonstrated high agreement with manual reference annotations (Dice 0.95 ± 0.02 for cortex and 0.90 ± 0.04 for medulla). Interreader agreement in 20 cases showed comparable overlap for cortex (Dice 0.97 ± 0.04) and greater variability for medulla (Dice 0.90 ± 0.11). A representative automated CT segmentation is shown in Figure 2. Quality control indicated sufficient model performance in 461 cases (89.9%), which comprised the final study cohort.

**Figure 2.**
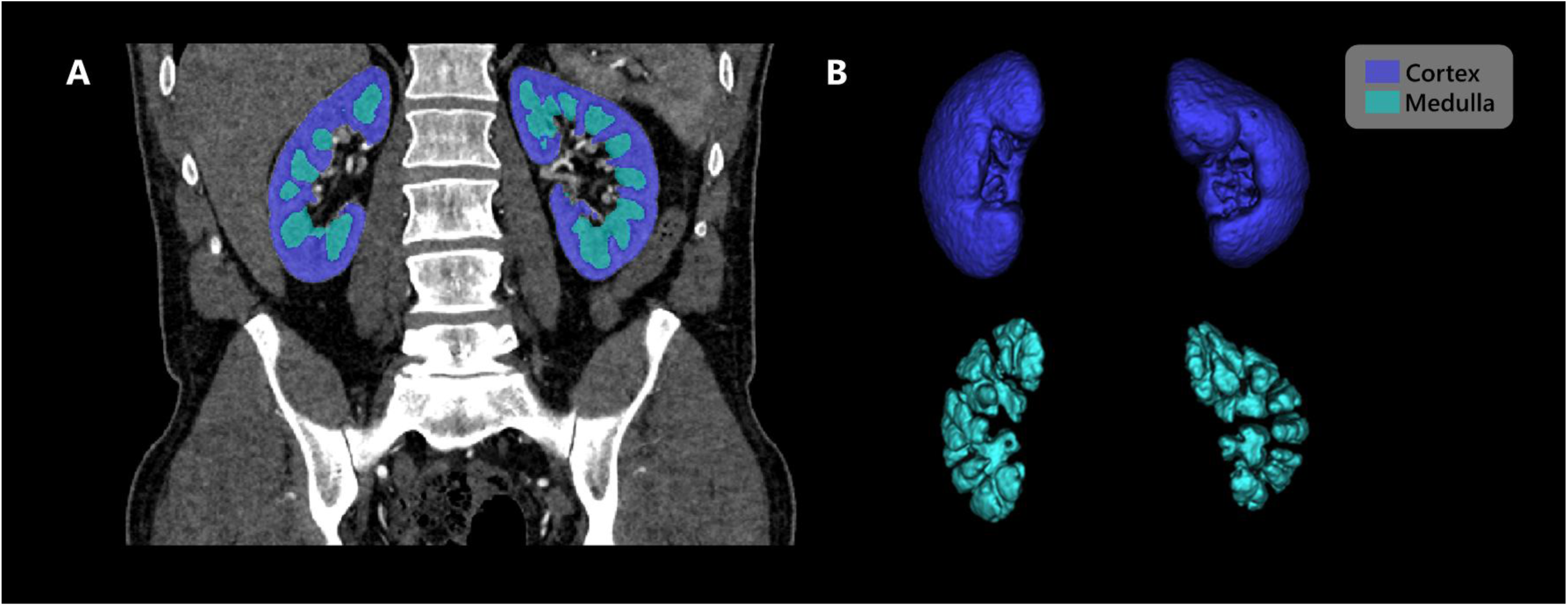
Representative example of fully automated arterial-phase CT segmentation. (A) Coronal CT image with automated cortical and medullary masks overlaid. (B) Three-dimensional renderings of the segmented cortical and medullary compartments.

**Figure 3.**
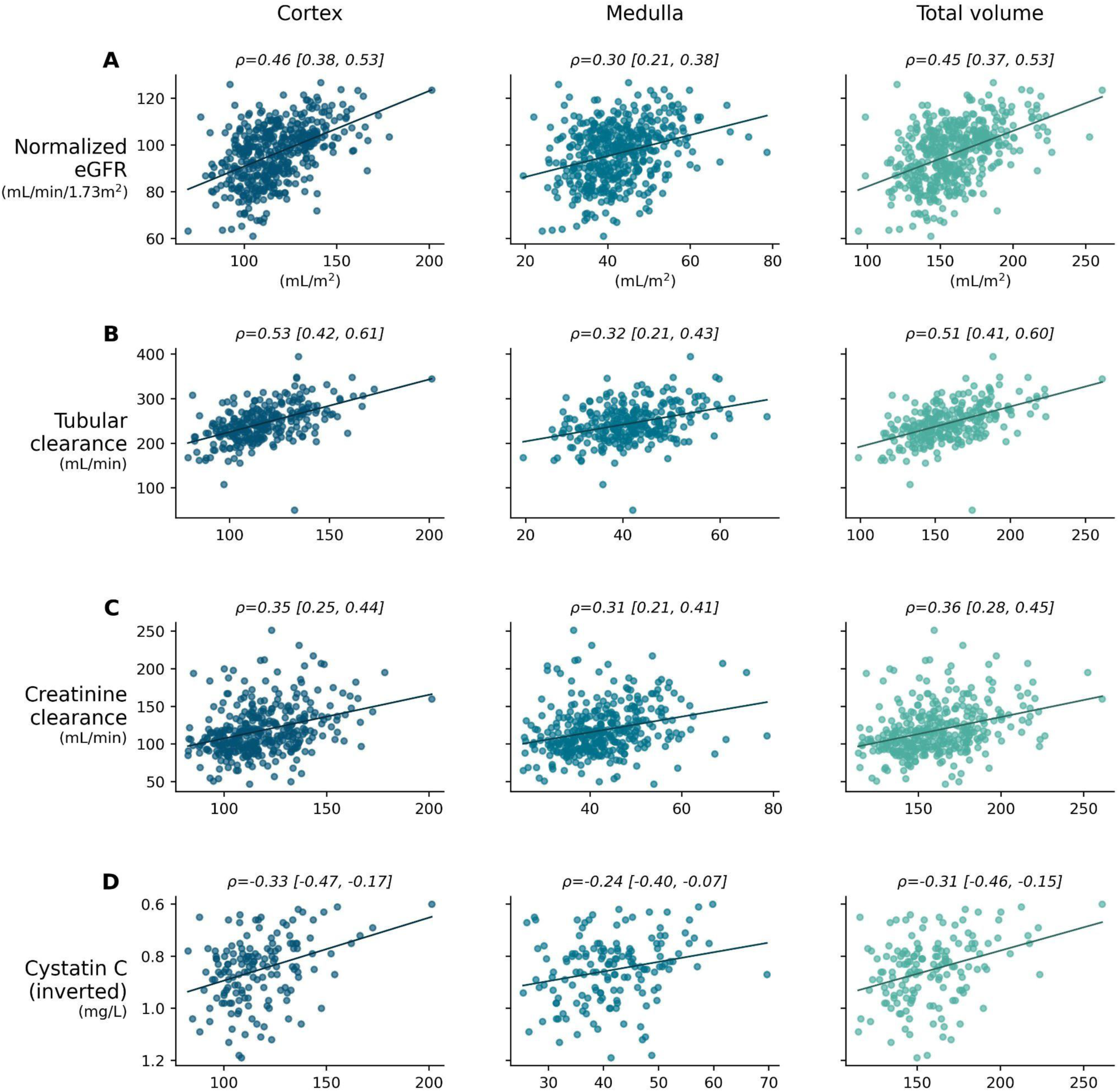
Scatter plots illustrating associations between global renal function parameters and BSA-normalized renal volumes. Columns represent cortical volume (left), medullary volume (middle), and total parenchymal volume (right). Spearman correlation coefficients (ρ) are shown for each association, with bootstrapped 95% confidence intervals in brackets. Serum cystatin C values are inverted to align correlation direction for visual comparison.

**Figure 4.**
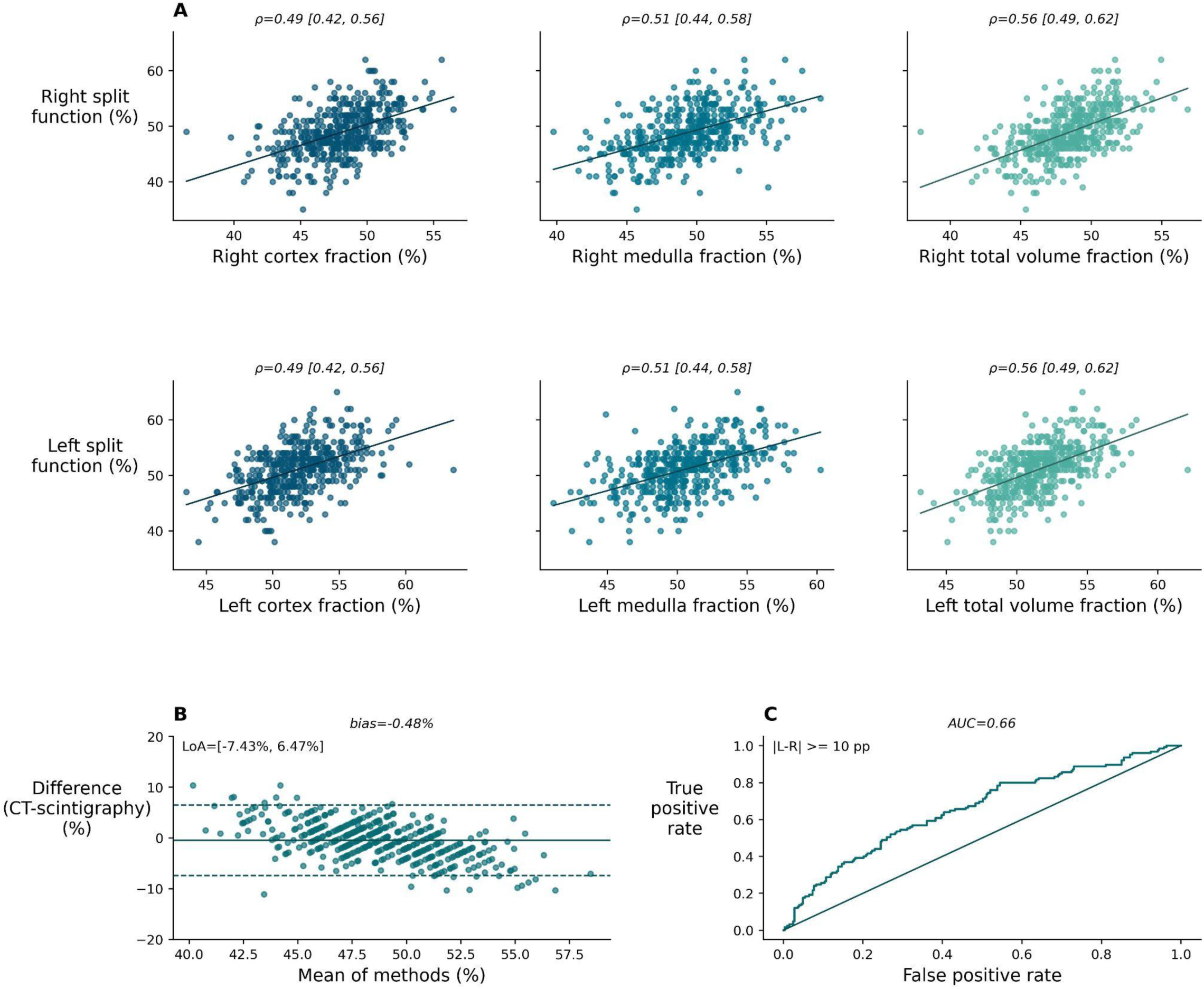
Agreement and discrimination of CT-derived renal volume fractions for split renal function. **(A)** Scatter plots showing correlations between CT-derived side-specific volume fractions and scintigraphy-derived split renal function; Spearman ρ values with bootstrapped 95% confidence intervals are shown. **(B)** Bland-Altman plot comparing CT- and scintigraphy-derived right renal fraction per donor (left fraction is complementary, 100 - right); solid lines indicate mean bias and dashed lines the 95% limits of agreement (LoA). **(C)** Receiver operating characteristic (ROC) curve for detection of clinically relevant split asymmetry (absolute difference ≥10 percentage points), with area under the curve (AUC) shown.

Mean BSA-normalized total parenchymal volume was 159.0 ± 24.1 mL/m², comprising cortical volume of 116.8 ± 18.2 mL/m² and medullary volume of 42.2 ± 8.4 mL/m². The mean cortical fraction (cortex/total parenchymal volume) was 0.73 ± 0.03.

### Associations Between Global Renal Function and Subcompartment Volumes

Using BSA-normalized volumes, cortical and total parenchymal volumes demonstrated moderate positive associations with eGFR (ρ = 0.46 and 0.45, respectively; both p < 0.001), whereas medullary volume showed a weaker association (ρ = 0.30; p < 0.001).

A similar pattern was observed across other global function measures, with cortical volume showing the strongest associations, closely followed by total parenchymal volume, and weaker correlations for medullary volume (Table 2).

**Table 2.** Associations between BSA-normalized CT-derived renal volumes and global renal function measures. Values are Spearman correlation coefficients (ρ), bootstrapped 95% confidence intervals are shown in brackets. Asterisks denote statistical significance (* p ≤ 0.01; ** p < 0.001). Negative correlations with cystatin C reflect inverse relationships with kidney function.

| CT-derived normalized volume | eGFR | 24-h creatinine clearance | Cystatin C | Scintigraphy tubular clearance |
| --- | --- | --- | --- | --- |
| Cortical volume | 0.46<br>[0.39, 0.53]** | 0.35<br>[0.25, 0.44]** | -0.33<br>[-0.47, -0.16]* | 0.53<br>[0.43, 0.62]** |
| Medullary volume | 0.30<br>[0.21, 0.38]** | 0.32<br>[0.22, 0.41]** | -0.24<br>[-0.39, -0.07]* | 0.32<br>[0.20, 0.43]** |
| Total parenchymal volume | 0.45<br>[0.37, 0.53]** | 0.37<br>[0.27, 0.45]** | -0.31<br>[-0.46, -0.15]* | 0.51<br>[0.41, 0.60]** |

In sensitivity analyses using absolute volumes and absolute eGFR (mL/min), associations were stronger, particularly for cortical volume (ρ = 0.69) and total parenchymal volume (ρ = 0.67), with a moderate association for medullary volume (ρ = 0.49) (all p < 0.001; n = 461).

### Split Renal Function: Correlation and Agreement

CT-derived side-specific volume fractions correlated strongly with scintigraphy-derived split renal function (n = 452), with ρ = 0.56 for total parenchymal volume fraction, 0.49 for cortical fraction, and 0.51 for medullary fraction (all p < 0.001).

Bland-Altman analysis demonstrated minimal mean bias between CT- and scintigraphy-derived left fractions (≤1 percentage point across compartments), with 95% limits of agreement of approximately ±7-8 percentage points and a mean absolute error of approximately 3 percentage points. Proportional bias was present for all compartments (slopes −0.65, −0.56, and −0.41; all p < 0.001), indicating compression of extreme asymmetry (Table 3).

**Table 3.** Relationship between BSA-normalized CT-derived volume fractions and scintigraphy split renal function. **(A)** Spearman correlations (ρ) between CT-derived side-specific volume fractions and scintigraphy split renal function, with bootstrapped 95% confidence intervals in brackets. **(B)** Bland-Altman analysis of CT versus scintigraphy right-sided fractions (CT minus scintigraphy; bias and 95% limits of agreement shown). **(C)** Diagnostic performance for detection of clinically relevant split asymmetry (absolute left-right difference ≥10 percentage points), expressed as area under the ROC curve (AUC). Asterisks indicate statistical significance (* p ≤ 0.01; ** p < 0.001 ).

**A) Correlation with scintigraphy split renal function**
| CT-derived fraction (%) | Left split function ($\rho$) | Right split function ($\rho$) | n |
| --- | --- | --- | --- |
| TKV fraction | 0.56 [0.49, 0.63]** | 0.56 [0.49, 0.63]** | 452 |
| Cortex fraction | 0.49 [0.42, 0.56]** | 0.49 [0.42, 0.56]** | 452 |
| Medulla fraction | 0.51 [0.44, 0.58]** | 0.51 [0.44, 0.58]** | 452 |

**B) Bland-Altman agreement for CT-derived vs scintigraphy-derived split fractions (right kidney)**
| Metric | Bias (%) | 95% limits of agreement (%) | Mean absolute error (%) | Proportional bias slope (%) | n |
| --- | --- | --- | --- | --- | --- |
| TKV fraction | -0.48 | -7.43 to 6.47 | 2.82 | -0.64** | 452 |
| Cortex fraction | -0.84 | -8.24 to 6.57 | 3.04 | -0.56** | 452 |
| Medulla fraction | 0.52 | -6.96 to 7.99 | 3.03 | -0.42** | 452 |

**C) Clinically relevant split asymmetry (scintigraphy absolute left-right difference $\geq 10$ pp)**
| Outcome | Value |
| --- | --- |
| Prevalence | 125/452 (27.7%) |
| AUC (TKV asymmetry from fractions) | 0.66 (n = 452; events = 125) |
| AUC (cortex asymmetry from fractions) | 0.62 (n = 452; events = 125) |
| AUC (medulla asymmetry from fractions) | 0.60 (n = 452; events = 125) |

### Clinically Relevant Split Asymmetry

Clinically relevant split asymmetry (absolute left-right difference ≥10 percentage points on scintigraphy) was present in 125 of 452 candidates (27.7%), including 44 of 172 male (25.6%) and 81 of 280 female candidates (28.9%). In the paired CT-scintigraphy subset (n = 452), CT-derived volume fractions demonstrated modest discriminative performance for detecting this threshold, with AUCs of 0.66 for total parenchymal volume fraction, 0.62 for cortical fraction, and 0.60 for medullary fraction (Table 3).

## Discussion

In this retrospective cohort of living kidney donor candidates, automated arterial-phase CT segmentation enabled reliable extraction of cortical, medullary, and total parenchymal volumes and demonstrated consistent associations with both global and split renal function. Cortical volume and total parenchymal volume correlated similarly with eGFR (ρ 0.46 and 0.45; p < 0.001), whereas medullary volume showed a weaker association (ρ 0.30). Similar patterns were observed for tubular clearance, 24-hour creatinine clearance, and cystatin C. These findings are physiologically coherent, as the cortex contains the majority of glomeruli and most directly reflects filtration capacity, whereas the medulla represents tubular architecture (14).

The trained segmentation model operates automatically on routine arterial-phase CT and achieved agreement with manual annotations comparable to human interreader variability (Dice 0.95 for cortex and 0.90 for medulla; interreader Dice 0.97 and 0.90, respectively). These findings are consistent with prior multisite validation studies (15). By eliminating manual and density-threshold-based segmentation, this automated approach reduces reader dependence and sensitivity to fixed Hounsfield unit thresholds (8,16), supporting more standardized implementation in clinical workflows.

Despite the narrow physiologic bandwidth of this healthy cohort (mean eGFR 96.3 ± 12.4 mL/min/1.73 m²; cortex/total parenchymal volume ratio 0.73 ± 0.03), cortical volume performed comparably to total parenchymal volume and maintained robust structure-function relationships. The limited variability in parenchymal composition and function inherently constrains incremental separation in donors; nevertheless, the preserved association between cortical volume and renal function supports the biologic relevance of cortical volumetry. Prior work has demonstrated age-related cortical decline paralleling GFR reduction (17), and large-scale automated segmentation studies have linked compartment volumes to kidney health phenotypes in broader populations, with effect sizes comparable to those observed in this healthy donor cohort (18). Greater incremental value of compartment-resolved metrics is therefore expected in older or CKD-enriched cohorts with more pronounced cortical thinning.

Regarding split function, CT-derived side-specific volume fractions correlated strongly with scintigraphic split renal function (ρ 0.49–0.56), with minimal mean bias but moderate individual variability. Discrimination for a ≥10-percentage-point asymmetry threshold was modest (AUC 0.60–0.65), despite the observed continuous correlation, likely because dichotomizing a continuous relationship reduces discrimination near the threshold (8). Because scintigraphic split-function estimates are subject to measurement variability (8,19,20), and the >=10-percentage-point cutoff is clinically pragmatic rather than uniformly outcome-validated (2,8), CT volumetry may be best interpreted as a complementary continuous marker that provides anatomically grounded context, particularly in borderline cases. Building on prior work linking CT volumetry to post-nephrectomy renal outcomes (4), future prospective studies should test whether CT-derived biomarkers enable improved outcome prediction and data-driven risk stratification. Beyond static volumetry, CT-based pipelines may also enable multiphase compartment-resolved attenuation analyses across arterial, portal venous, and excretory phases to further refine functional characterization.

This study is limited by its retrospective, single-center design. The long inclusion period introduces protocol- and scanner-related variability. Global renal function was primarily assessed using serum creatinine-based eGFR, which is less precise at high-normal kidney function and may attenuate observed correlations in this screened donor population. Although additional metrics (24-hour creatinine clearance, serum cystatin C, and scintigraphy-derived tubular clearance) were available in subsets, a single measured reference standard was not available for all participants. In addition, the donor population represents a restricted clinical spectrum with limited cortical thinning and functional dispersion, which may underestimate the incremental value of subcompartment analysis.

In conclusion, automated arterial-phase CT segmentation provides reproducible, physiologically interpretable volumetric biomarkers from imaging already embedded in donor workup. In healthy donors, cortical volume performs comparably to total parenchymal volume for global function assessment, and CT-derived side fractions track scintigraphic split function at the cohort level. Prospective multicenter validation in more heterogeneous populations is warranted. At present, automated volumetry may serve as a complementary metric to identify clinically relevant compartmental and side asymmetries during routine donor evaluation.

## Data Availability

The de-identified data that support the findings of this study are available from the corresponding author upon reasonable request, subject to institutional and data protection regulations.

## Abbreviations

AI: Artificial Intelligence
BSA: Body Surface Area
CT: Computed Tomography
Dice: Dice Similarity Coefficient
eGFR: Estimated Glomerular Filtration Rate

## References

1. Sebastià C, Peri L, Salvador R, et al. Multidetector CT of Living Renal Donors: Lessons Learned from Surgeons. RadioGraphics. Radiological Society of North America; 2010;30(7):1875–1890. doi: 10.1148/rg.307105032.

2. Lentine KL, Kasiske BL, Levey AS, et al. KDIGO Clinical Practice Guideline on the Evaluation and Care of Living Kidney Donors. Transplantation. 2017;101(8S):S7. doi: 10.1097/TP.0000000000001769.

3. Poggio ED, Hila S, Stephany B, et al. Donor Kidney Volume and Outcomes Following Live Donor Kidney Transplantation. American Journal of Transplantation. Elsevier; 2006;6(3):616–624. doi: 10.1111/j.1600-6143.2005.01225.x.

4. Eum SH, Lee H, Ko EJ, Cho HJ, Yang CW, Chung BH. Comparison of CT volumetry versus nuclear renography for predicting remaining kidney function after uninephrectomy in living kidney donors. Sci Rep. Nature Publishing Group; 2022;12(1):5144. doi: 10.1038/s41598-022-09187-9.

5. Denic A, Alexander MP, Kaushik V, et al. Detection and Clinical Patterns of Nephron Hypertrophy and Nephrosclerosis Among Apparently Healthy Adults. American Journal of Kidney Diseases. Elsevier; 2016;68(1):58–67. doi: 10.1053/j.ajkd.2015.12.029.

6. Muto NS, Kamishima T, Harris AA, et al. Renal cortical volume measured using automatic contouring software for computed tomography and its relationship with BMI, age and renal function. European Journal of Radiology. Elsevier; 2011;78(1):151 –156. doi: 10.1016/j.ejrad.2009.10.005.

7. Wahba R, Franke M, Hellmich M, et al. Computed Tomography Volumetry in Preoperative Living Kidney Donor Assessment for Prediction of Split Renal Function. Transplantation. 2016;100(6):1270. doi: 10.1097/TP.0000000000000889.

8. López-Abad A, Prudhomme T, Pecoraro A, et al. Can CT or MRI volumetry substitute scintigraphy in living kidney donor evaluation? A systematic review. World J Urol. 2024;42(1):382. doi: 10.1007/s00345-024-05024-y.

9. Reisert M, Russe M, Elsheikh S, Kellner E, Skibbe H. Deep Neural Patchworks: Coping with Large Segmentation Tasks. 2022; doi: 10.48550/arXiv.2206.03210.

10. Reisert M, Russe M, Elsheikh S, Kellner E, Skibbe H. Deep Neural Patchworks: Coping with Large Segmentation Tasks. arXiv.org. 2022. https://arxiv.org/abs/2206.03210v1. Accessed February 21, 2026.

11. Nora - The Medical Imaging Platform. . https://www.nora-imaging.com/. Accessed September 1, 2023.

12. Mosteller RD. Simplified calculation of body-surface area. N Engl J Med. 1987 Oct 22;317(17):1098. doi: 10.1056/NEJM198710223171717.

13. Inker LA, Eneanya ND, Coresh J, et al. New Creatinine- and Cystatin C–Based Equations to Estimate GFR without Race. New England Journal of Medicine. Massachusetts Medical Society; 2021;385(19):1737–1749. doi: 10.1056/NEJMoa2102953.

14. Kuhnel L, Vu T, Wood ST, Francis RS, Trnka P, Ellis RJ. Relationship between cortical and medullary thickness and glomerular filtration rate among living kidney donors. Internal Medicine Journal. 2023;53(3):431–435. doi: 10.1111/imj.16033.

15. Korfiatis P, Denic A, Edwards ME, et al. Automated Segmentation of Kidney Cortex and Medulla in CT Images: A Multisite Evaluation Study. Journal of the American Society of Nephrology. 2022;33(2):420. doi: 10.1681/ASN.2021030404.

16. Gardan E, Jacquemont L, Perret C, et al. Renal cortical volume: High correlation with pre- and post-operative renal function in living kidney donors. European Journal of Radiology. Elsevier; 2018;99:118–123. doi: 10.1016/j.ejrad.2017.12.013.

17. Wang X, Vrtiska TJ, Avula RT, et al. Age, kidney function, and risk factors associate differently with cortical and medullary volumes of the kidney. Kidney International. Elsevier; 2014;85(3):677–685. doi: 10.1038/ki.2013.359.

18. Kellner E, Sekula P, Lipovsek J, et al. Imaging Markers Derived From MRI-Based Automated Kidney Segmentation—an Analysis of Data From the German National Cohort (NAKO Gesundheitsstudie). Dtsch Arztebl Int. 2024;(Forthcoming):arztebl.m2024.0040. doi: 10.3238/arztebl.m2024.0040.

19. Taylor AT, Brandon DC, de Palma D, et al. SNMMI Procedure Standard/EANM Practice Guideline for Diuretic Renal Scintigraphy in Adults With Suspected Upper Urinary Tract Obstruction 1.0. Seminars in Nuclear Medicine. 2018;48(4):377–390. doi: 10.1053/j.semnuclmed.2018.02.010.

20. Brink A, Libhaber E, Levin M. Renogram image characteristics and the reproducibility of differential renal function measurement. Nuclear Medicine Communications. 2021;42(8):866. doi: 10.1097/MNM.0000000000001408.

